# Three montages for Transcranial Electric Stimulation in predicting the early post-surgery outcome of the facial nerve functioning

**DOI:** 10.1101/2024.02.28.24302993

**Authors:** Mikael Gian Andrea Izzo, Davide Rossi Sebastiano, Valentina Catanzaro, Ylenia Melillo, Ramona Togni, Elisa Visani, Jacopo Falco, Cecilia Casali, Marco Gemma, Paolo Ferroli, Annamaria Gallone, Cazzato Daniele, Grazia Devigili, Sara Alverà, Paola Lanteri

## Abstract

**Objective:** We assessed the Transcranial Electrical Stimulation (TES)-induced Corticobulbar-Motor Evoked Potentials (Cb-MEPs) evoked from Orbicularis Oculi (Oc) and Orbicularis Oris (Or) muscles with FCC5h/FCC6h-Mz, C3/C4-Cz and C5/C6/-Cz stimulation, during IntraOperative NeuroMonitoring (IONM) in 30 patients who underwent skull-base surgery.

**Methods:** before (T0) and after (T1) the surgery, we compared the peak-to-peak amplitudes of Cb-MEPs obtained from TES with C3/C4-Cz, C5/C6-Cz and FCC5h/FCC6h-Mz. Then, we compared the response category (present, absent and peripheral) related to different montages. Finally, we classified the Cb-MEPs data from each patient for concordance with clinical outcome and we assessed the diagnostic measures for Cb-MEPs data obtained from FCC5h/FCC6h-Mz, C3/C4-Cz and C5/C6-Cz TES stimulation.

**Results:** Both at T0 and T1, FCC5h/FCC6h-Mz stimulation evoked larger Cb-MEPs than C3/C4-Cz, less peripheral responses from direct activation of facial nerve than C5/C6-Cz. FCC5h/FCC6h-Mz stimulation showed the best accuracy and specificity of Cb-MEPs for clinical outcomes.

**Conclusions:** FCC5h/FCC6h-Mz stimulation showed the best performances for monitoring the facial nerve functioning, maintaining excellent diagnostic measures even at low stimulus voltages. <u>Significance:</u> We demonstrated that FCC5h/FCC6h-Mz TES montage for Cb-MEPs in IONM has good accuracy in predicting the post-surgery outcome of facial nerve functioning.

**Highlights:**

- At low voltage, FCC5h/FCC6h-Mz stimulation evoked larger Cb-MEPs than C3/C4-Cz
- At low voltage, FCC5h/FCC6h-Mz stimulation induced less peripheral responses by direct activation of facial nerve than C5/C6-Cz
- FCC5h/FCC6h-Mz stimulation is safe and increases the accuracy and the specificity of Cb-MEPs with respect to facial nerve outcome

## 1. Introduction

IntraOperative NeuroMonitoring (IONM) of cranial nerves, especially of the facial nerve (Chang et al., 1999), is important to spare their functioning and to reduce the incidence of new neurological deficit in several neurosurgical procedures, above all in brainstem and skull base surgery (Isackson et al., 2005; Morota et al., 2010; Kodama et al., 2022).

Continuous electromyographic (EMG) monitoring of muscles innervated by cranial nerves for identification of type A spontaneous activity (Romstöck et al., 2000), blink-reflex (Deletis et al., 2009a), brainstem auditive evoked and acoustic nerve action potentials (James and Husain, 2005; Youssef and Downes, 2009), are routinely used in these surgical procedures as long as the direct stimulation of cranial nerves or of cranial nerve nuclei in the brainstem (Dickins and Graham, 1991; Lin et al., 2006; Fernández-Conejero et al., 2022).

Among other techniques, the functioning of cranial nerves can be monitored by Transcranial Electrical Stimulation (TES)-induced Corticobulbar-Motor Evoked Potentials (Cb-MEPs) in their innervated muscles (Dong et al., 2005, Deletis et al., 2009b, Deletis et al., 2011). Since Cb-MEPs are the only technique able to provide information on the functioning of the whole motor pathway, from cortical motor areas through cortico-bulbar tract and from brainstem motor nuclei to cranial nerves, their use in IONM of skull-base surgery is growing in importance. Nevertheless, a global consensus on some technical specifications is lacking, and some methodological points are still disputed.

Since there is a wide anatomic (and thus functional) variability in the individual location of the motor cortical area of the human brain (Eichert et al., 2021), a presurgical brain mapping of motor functions should be performed with functional neuroimaging techniques or transcranial magnetic stimulation (for recent reviews on topic see Sollmann et al, 2021 and Voets et al., 2022), thus improving the preoperative surgical plan. The presurgical assessment of cortical motor areas is an open issue of the utmost relevance for the surgery involving supratentorial regions of the brain (Rammelloo et al., 2023), where sparing the eloquent cortical areas is mandatory, but it would be important as well in the brainstem and skull base surgery, helping in the customization of a most reliable TES montage in each patient. However, it is not always easy to perform presurgical brain mapping, therefore, in the daily practice, the individual anatomical landmarks and skull measure are used to place electrodes for TES in a “standard” way.

Most of the groups usually perform TES with electrodes placed in C3-C4/C4-C3 or C3/C4-Cz (Zhou and Kelly, 2001; Akagami et al., 2005; Dong et al., 2005, Deletis et al., 2009b, Deletis et al., 2011), or in a similar way (Goto et al., 2010). On the other hand, Cb-MEPs of similar amplitude can be obtained (with even lower stimulus voltage) with C5/C6-Cz montage (Verst et al., 2012; Verst et al., 2013).

Although C3/C4-Cz is a widely used montage, C3/C4 are generally considered skull points centred on the sensory-motor hand areas (Shibasaki H., 2008; Goto et al., 2010; Holmes et al, 2019); on the contrary, the C5/C6-Cz montage was only used by a single group (Verst et al., 2012; Verst et al., 2013), and confirmative studies are needed before its extensive use in the daily routine, even considering that the Authors did not assess the Cb-MEP results with the post-surgical outcome of the facial nerves. Moreover, during neuronavigation, it is common to see that C5 and C6 ideally project in areas lying just behind the precentral gyrus, in the somatosensory areas, due to the peculiar inclination of the Rolandic sulcus. Hence, neither C3/C4-Cz nor C5/C6-Cz montage probably ensure the best possible localization of electrodes, in order to obtain Cb-MEPs with a very focal stimulus.

In our experience, a good solution to evocate Cb-MEP is the use of FCC5h/FCC6h-Mz stimulation, where Mz is located 1 cm anteriorly to Cz, while FCC5h and FCC6h are placed as suggested by the international electrode system for high-resolution EEG (Oostenveld and Praamstra, 2001); in the vast majority of the patients FCC5h and FCC6h lie anteriorly to the Rolandic sulcus, in a very favourable location for a stimulation focused on the mouth motor area; nevertheless, to our knowledge, there are no works in the literature in which this type of montage was used.

In this study, we aimed to assess the safety and the feasibility of the TES-induced Cb-MEPs evoked from FCC5h/FCC6h-Mz montage in a group of patients who underwent brainstem or skull-base surgery. Moreover, we want to compare the amplitude and the accuracy of Cb-MEPs evoked with this montage (FCC5h/FCC6h-Mz), with respect to Cb-MEPs evoked with a C3/C4-Cz and a C5/C6/-Cz ones.

## 2. Methods

### 2.1. Study population and clinical parameters

We enrolled 30 consecutive adult patients (14 females and 16 males; age ranging from 18.1 to 73.0 years, mean 47.9 + 15.0) who underwent brainstem or skull-base surgery with IONM at the Neurological Institute “Carlo Besta” of Milan, from January, 1^st^, 2023 to June, 30^th^, 2023.

One week before and one week after the surgery, an expert neurologist (MI) performed the six-point grading scale of House-Brackmann (HB, House and Brackmann, 1985) of each patient to assess the functioning of the facial nerves. Patients bearing clinical disorders such as neurological, psychiatric or systemic major diseases were excluded from the study, as well as patients with preoperative moderately severe, severe or global dysfunction of at least one facial nerve (HB = IV, V or VI).

The study was approved by the local ethics committee (study “Indicators of outcome, disability and quality of life in neurosurgery”, number 46/17); all the patients signed their informed consent.

### 2.2. Anesthesiological parameters

In all of the patients total intravenous anesthesia (TIVA) was induced and maintained with propofol and remifentanil administered by Target Controlled Infusion (TCI). The depth of anesthesia was continuously monitored throughout surgery by means of the Bispectral Index (BIS). The propofol TCI targeted a BIS value ranging from 40 to 60 (Avidan et al., 2008).

Neuromuscular blockade was limited to a single 0.6 mg/Kg i.v. dose of rocuronium (a non-depolarizing agent) administered after induction of anesthesia to facilitate tracheal intubation. Cb-MEPs recording started only after complete recovery of the train-of-four stimulation. I.v. sugammadex was administered to completely reverse neuromuscular blockade if needed. Intraoperative standard anesthesiological monitoring was applied.

### 2.3. Introperative neuromonitoring

IONM was performed with NIM Eclipse (Medtronic, Italy); corkscrew needles were used for TES, and two monopolar non-teflon subdermal needle electrodes were used for recording TES-induced Cb-MEPs and spontaneous activity of the Orbicularis Oculi (Oc) and Orbicularis Oris (Or) muscles; 100 Hz-1KHz bandpass filters were applied. TES was performed with a short train of 5 stimuli of 0.5 ms duration with inter-stimulus interval of 2 ms, followed by a single stimulus of 0.5 ms duration at 90 ms after the train; the single stimulus was performed to disentangle between “central” and “peripheral” responses (Deletis et al., 2009b). In each patient, TES was set at the voltage sufficient to obtain from FCC5h/FCC6h-Mz the maximal peak-to-peak amplitude of Cb-MEPs induced by the train of 5 stimuli without any response greater than 10 μV induced by the single stimulus, in at least one of the two muscles explored (Oc and Or).

When it was impossible to evoke a Cb-MEPs without a peripheral activation, the voltage was set at the lowest value at which the peripheral response reached a peak-to-peak amplitude of at least 10 μV (this method was applied only for patient #12).

For the subsequent analysis, we collected the peak-to-peak amplitude of Cb-MEPs from Oc and Or muscles of the affected side obtained by TES with C3/C4-Cz, C5/C6-Cz and FCC5h/FCC6h-Mz montage, just before the craniotomy (baseline, T0) and immediately after the end of neurosurgery (T1). The same voltage of TES determined for FCC5h/FCC6h-Mz stimulation was applied for C3/C4-Cz and C5/C6-Cz ones. Again, the same voltage was applied at T0 and T1.

Then, on the basis of the peak-to-peak amplitudes, the responses were converted to the following categorical data:

- PRESENT: peak-to-peak amplitude greater than 10 μV induced by the train of stimuli with peak-to-peak amplitude smaller than 10 μV induced by single stimulus.
- ABSENT: peak-to-peak amplitude ranging from 0 to 10 μV induced by both the train of stimuli and single stimulus.
- PERIPHERAL: peak-to-peak amplitude greater than 10 μV induced by single stimulus (independently by the peak-to-peak amplitude induced by the train of stimuli). Cb-MEPs with response greater than 10 μV induced by single stimulus were considered polluted by the direct stimulation of the facial nerve.

During the surgical procedures (i.e. after the T0 and before the T1 measures) IONM proceeded as usually performed in our Institute for clinical and diagnostic purpose, i.e. by considering only the Cb-MEPs obtained from the standard C3/C4-Cz montage.

### 2.4. Statistical analysis

#### 2.4.1. Comparison between the peak-to-peak amplitudes of the Cb-MEPs

To evaluate differences in the responses related to different point of stimulation, the peak-to-peak amplitudes of the Cb-MEPs obtained from TES with C3/C4-Cz, C5/C6-Cz and FCC5h/FCC6h-Mz montages were compared separately for Oc and Or muscles and for T0 and T1. Since the data were not normally distributed, Friedman test was applied to compare the values. Then, Wilcoxon signed-rank test with Bonferroni correction for multiple comparisonwas used as the post hoc test and to compare each amplitude between T0 and T1. Finally, to evaluate the response category (present, absent or peripheral) related to different stimulation montages, a χ^2^ test was used for each muscles (Oc and Or) and Time (T0 and T1).

#### 2.4.2. Comparison between IONM data and postsurgical outcome: calculation of diagnostic measures

The concordance or discordance between Cb-MEPs variation and the surgical outcome in term of functioning of facial nerves was assessed separately for each patient and for each montage used as follow.

Firstly, a decrease of at least 50% in the peak-to-peak amplitude in at least one of Oc and Or was considered as “positive” for a postsurgery worsening of facial nerve functioning, while the presence at T1 of Cb-MEPs with peak-to-peak amplitude ≥ 50% then those at T0 in both Oc and Or was considered as “negative” for a postsurgery worsening of facial nerve functioning. If at T0 and/or at T1 we obtained a peripheral response in both Oc and Or or if we did not evoke any Cb-MEP in both Oc and Or at T0, we considered as “not applicable” the outcome prediction on the basis of Cb-MEPs data.

Then, the clinical assessment of the facial nerves was taken as reference outcome, discriminating between good (unchanged HB) and poor (worsened HB) surgical outcome.

The Cb-MEPs data from each patient were classified with respect to concordance with clinical outcome as:

- TRUE NEGATIVE (TN): concordance of the Cb-MEP “negative” outcome prediction for a worsening of facial nerve functioning with unchanged HB scale.
- FALSE NEGATIVE (FN): concordance of the Cb-MEP “negative” outcome prediction for a worsening of facial nerve functioning with worsened HB scale.
- TRUE POSITIVE (TP): concordance of the Cb-MEP “positive” outcome prediction for a worsening of facial nerve functioning with worsened HB scale.
- FALSE POSITIVE (FP): concordance of the Cb-MEP “positive” outcome prediction for a worsening of facial nerve functioning with unchanged HB scale.
- NOT APPLICABLE (NA) outcome prediction, if Cb-MEPs data at T0 and T1 were not comparable.

Figure 1 graphically shows the method used to compare the prediction based on Cb-MEPs data with the clinical post-surgical outcome.

**Figure 1:**
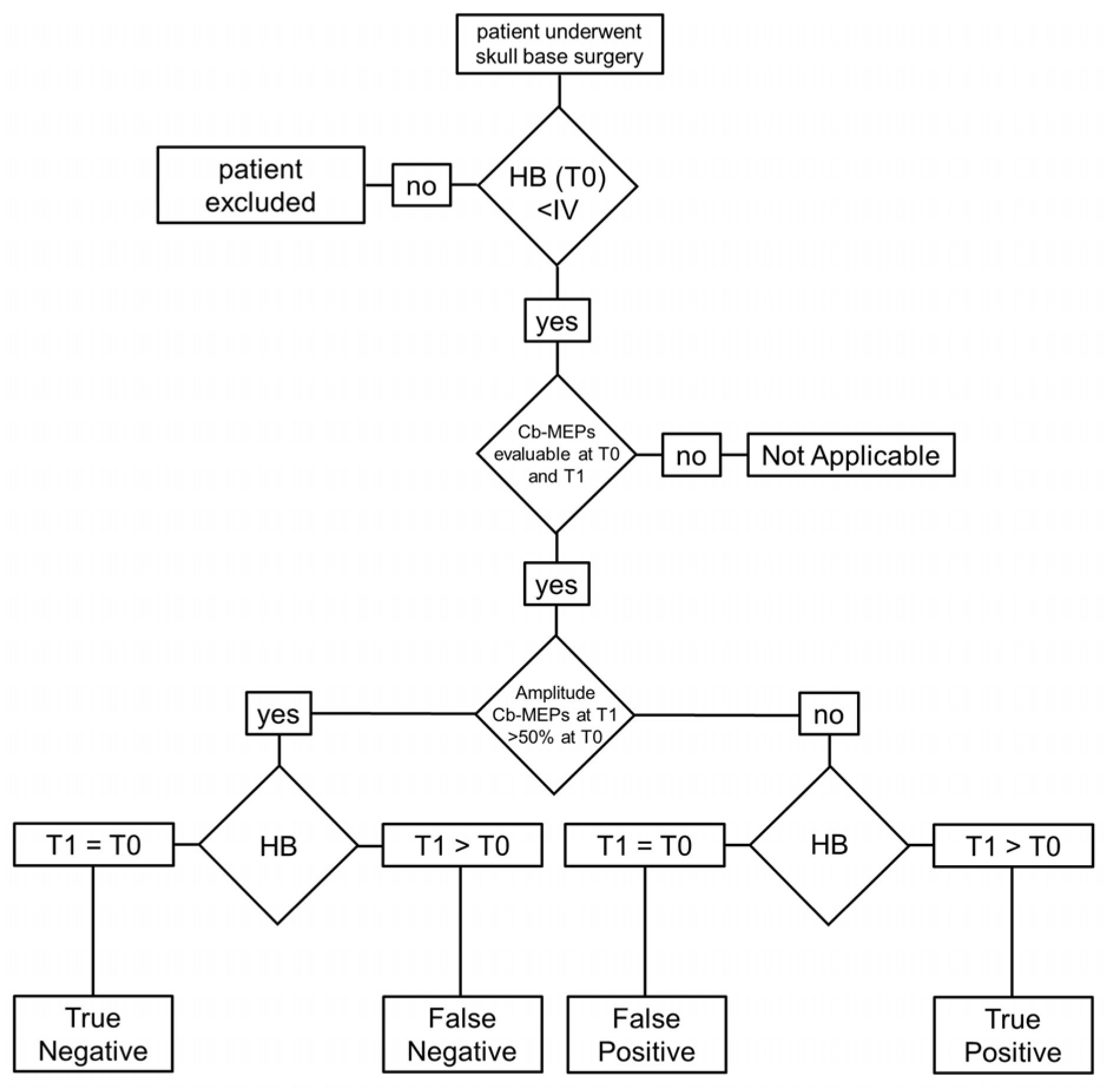
flowchart illustrating how the concordance between IONM data and postsurgical outcome was determined.

Finally, we calculated the following diagnostic measures:

- SENSITIVITY = TP/(TP+FN)
- SPECIFICITY = TN/(TN+FP)
- NEGATIVE PREDICTIVE VALUE (NPV) = TN/(TN+FN)
- POSITIVE PREDICTIVE VALUE (PPV) = TP/(TP+FP)
- ACCURACY = (TN+TP)/(TN+FN+TP+FP);

All the diagnostic measures were assessed for Cb-MEPs data obtained from FCC5h/FCC6h-Mz, C3/C4-Cz and C5/C6-Cz TES stimulation. Data were calculated with 95% Confidence Interval (CI), as suggested by the STARD initiative (Bossuyt et al., 2003).

## 3. Results

### 3.1. Clinical data

All patients included in the study had a tumor located in cerebello-pontine angle, nearby the fourth ventricle, posterior cranial fossa, anterior pons, cranio-cervical junction and spheno-petro-clival region in 17 (56.7%), 7 (23.3%), 3 (10.0%),1 (3.3%), 1 (3.3%) and 1 (3.3%) case(s), respectively. Acoustic neurinoma was the most represented tumor, since it was present in 10 (33.3%) patients, followed by meningioma which was present in 5 (16.7%) patients; the remaining patients showed lesions with various histology and different grading of malignancy.

Twenty-four (80.0%) patients did not show any abnormalities in the facial nerves functioning before surgery (HB = I), while 6 (20.0%) of them showed mild to moderately severe abnormalities (HB = II-IV). After surgery, worsening of the facial nerves functioning was observed in 9 (30.0%) patients, while in 21 (70.0%) HB score was unchanged.

Because of the presence of moderately severe abnormalities of the facial nerves functioning (HB scale = IV), 3 out of the 30 (10.0%) patients enrolled (#16, #20 and #26) were excluded from the statistical analysis, while the other 27 patients met the study’s inclusion criteria.

**Table 1** summarizes the clinical data for each patient.

**Table 1:**
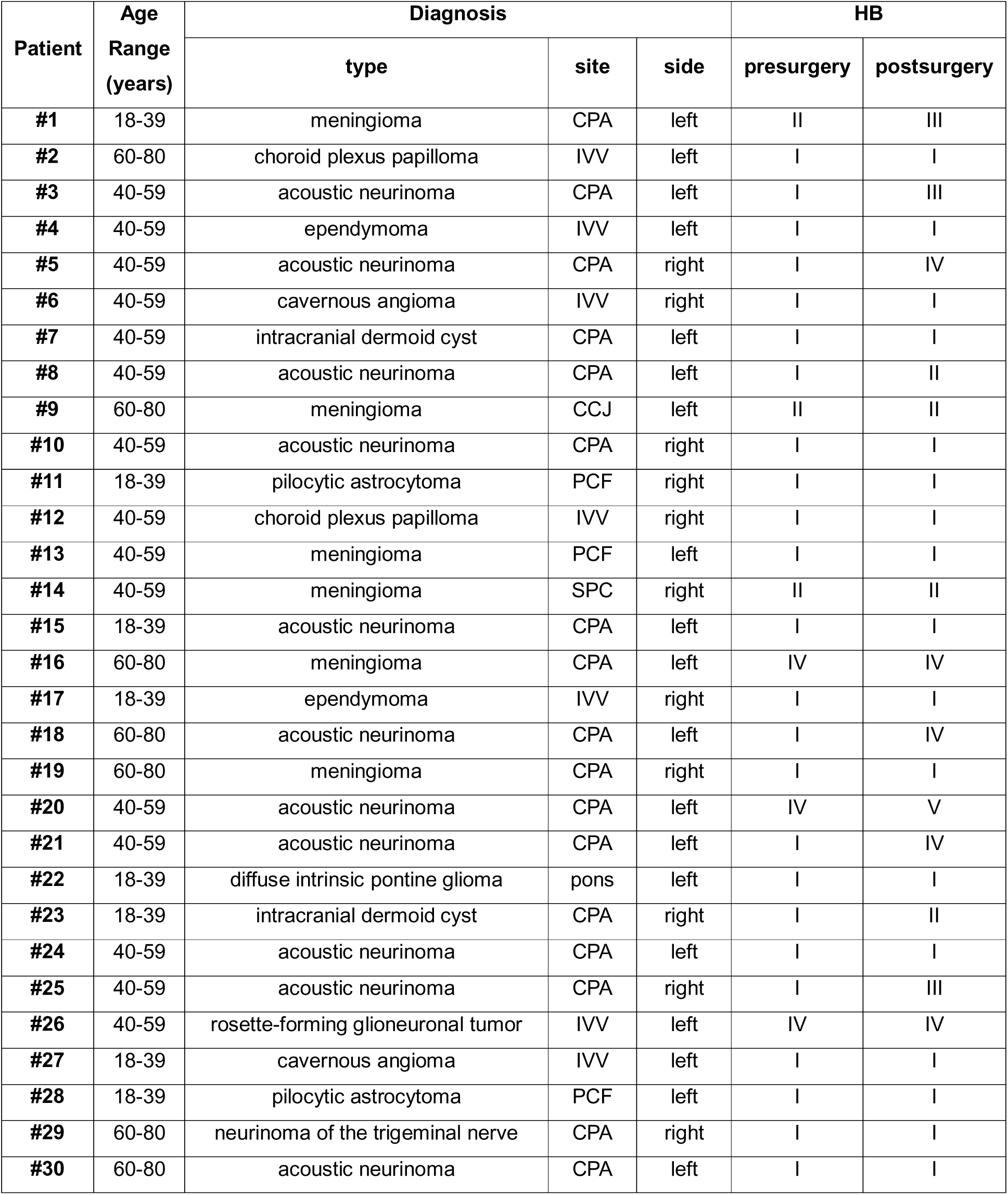
Clinical data. HB=House-Brackmann scale; CPA=cerebello-pontine angle; CCJ=cranio-cervical junction; IVV=IV ventricle; PCF=posterior cranial fossa; SPC= spheno-petro-clival region.

### 3.2. Anesthesiological parameters

Through appropriate propofol and remifentanil TCI adjustments the target BIS interval of 40-60 was always maintained and no clinically significant hemodynamic alteration occurred. Halogenated gas and nitrous oxide were never administered.

### 3.3. Introperative neuromonitoring and statistical analysis

It was possible to perform IONM throughout all the surgical procedures. No side effects and especially neither seizures nor after-discharges induced by TES were observed.

**Table 2** summarizes the IONM data for each patient; the response induced by the three different TES-montages were different both for the Oc and Or, at T0 and T1, while no significantly differences were found between T0 and T1 for all the responses of Oc and Or, considering each montage alone (see the upper part of **Table 3**).

**Table 2:** neurophysiological data (facial nerve) Ab= absent response; Cb-MEPs= Corticobulbar-Motor Evoked Potentials; PR=peripheral response.

| Patient | Stimulus intensity (microV) | Cb-MEPs amplitude (microV) |  |  |  |  |  |  |  |  |  |  |  |
| --- | --- | --- | --- | --- | --- | --- | --- | --- | --- | --- | --- | --- | --- |
|  |  | FCC5h/FCC6h-Mz |  |  |  | C5/C6-Cz |  |  |  | C3/C4-Cz |  |  |  |
|  |  | Orbicularis Oculi |  | Orbicularis Oris |  | Orbicularis Oculi |  | Orbicularis Oris |  | Orbicularis Oculi |  | Orbicularis Oris |  |
|  |  | T0 | T1 | T0 | T1 | T0 | T1 | T0 | T1 | T0 | T1 | T0 | T1 |
| #1 | 140 | 28 | PR | 120 | PR | 28 | PR | 60 | PR | Ab | Ab | Ab | Ab |
| #2 | 220 | 38 | 46 | PR | PR | 38 | PR | PR | PR | 30 | 128 | PR | PR |
| #3 | 120 | 20 | Ab | 440 | Ab | 25 | Ab | 154 | Ab | 15 | Ab | 110 | Ab |
| #4 | 190 | 701 | 590 | 812 | 867 | 721 | 375 | PR | PR | 280 | 375 | 202 | 333 |
| #5 | 170 | 50 | Ab | 148 | Ab | 25 | Ab | 131 | Ab | Ab | Ab | Ab | Ab |
| #6 | 200 | 50 | 158 | PR | PR | 22 | 92 | PR | PR | 27 | 43 | PR | PR |
| #7 | 140 | 59 | Ab | 153 | 114 | 31 | Ab | 123 | 92 | Ab | Ab | 194 | 53 |
| #8 | 110 | 208 | 144 | 166 | 279 | 228 | 117 | 52 | 35 | Ab | Ab | Ab | Ab |
| #9 | 120 | 34 | 75 | PR | PR | 39 | Ab | PR | PR | 41 | 57 | PR | PR |
| #10 | 270 | 522 | 533 | 153 | 279 | Ab | 385 | 131 | 179 | 239 | 408 | Ab | Ab |
| #11 | 120 | 82 | 44 | 658 | 300 | 45 | PR | 261 | PR | Ab | PR | Ab | PR |
| #12 | 230 | Ab | Ab | PR | PR | Ab | Ab | PR | PR | Ab | Ab | PR | PR |
| #13 | 140 | PR | 377 | PR | 138 | PR | 269 | 120 | 201 | 127 | 125 | Ab | Ab |
| #14 | 200 | 55 | 132 | 327 | 305 | 87 | 715 | 418 | 553 | Ab | 130 | 322 | 77 |
| #15 | 160 | 270 | 496 | 565 | 554 | Ab | Ab | Ab | Ab | Ab | Ab | Ab | Ab |
| #16 | Patient excluded |  |  |  |  |  |  |  |  |  |  |  |  |
| #17 | 150 | 95 | 119 | 214 | 353 | 57 | Ab | 54 | 87 | 53 | Ab | Ab | Ab |
| #18 | 150 | 70 | Ab | 135 | PR | Ab | Ab | 363 | 200 | Ab | Ab | 331 | PR |
| #19 | 290 | 425 | 504 | 494 | 538 | PR | PR | PR | PR | 227 | 210 | 363 | 201 |
| #20 | Patient excluded |  |  |  |  |  |  |  |  |  |  |  |  |
| #21 | 180 | 190 | PR | PR | PR | PR | PR | PR | PR | PR | PR | PR | PR |
| #22 | 130 | 52 | PR | 234 | 240 | Ab | Ab | 155 | 160 | Ab | Ab | Ab | Ab |
| #23 | 310 | Ab | Ab | 106 | PR | 48 | Ab | 135 | Ab | Ab | Ab | 331 | PR |
| #24 | 150 | 110 | 117 | 52 | 43 | 130 | PR | PR | PR | Ab | Ab | Ab | Ab |
| #25 | 180 | 57 | 43 | 211 | 235 | 58 | Ab | 210 | 140 | 63 | 60 | 216 | PR |
| #26 | Patient excluded |  |  |  |  |  |  |  |  |  |  |  |  |
| #27 | 230 | 285 | PR | PR | PR | PR | PR | PR | PR | 330 | PR | PR | PR |
| #28 | 170 | 53 | 47 | PR | PR | 28 | 26 | PR | PR | 54 | 52 | PR | PR |
| #29 | 200 | 212 | 245 | 458 | 482 | 125 | 103 | 556 | 290 | 194 | 204 | 151 | 275 |
| #30 | 170 | 69 | 45 | 124 | 108 | 41 | 30 | 152 | 86 | 54 | 28 | 96 | 18 |

**Table 3:** statistical analysis. N= number; Oc= Orbicularis Oculi; Or= Orbicularis Oris; NS= not significant

| Comparison of the peak-to-peak amplitudes of the Cb-MEPs at T0 and T1 |  |  |  |  |  |  |  |
| --- | --- | --- | --- | --- | --- | --- | --- |
| Muscle | Timing | N | Median (25<sup>th</sup>-75<sup>th</sup> percentile) amplitude (μV) |  |  | χ<sup>2</sup> (2) | p |
|  |  |  | FCC5h/FCC6h-Mz | C5/C6-Cz | C3/C4-Cz |  |  |
| Oc | T0 | 23 | 57 (38-110) | 38 (22-58) | 0 (0-53) | 12.51 | 0.002 |
|  | T1 | 18 | 61 (0-180) | 13 (0-155) | 36 (0-126) | 10.61 | 0.005 |
|  | Z |  | -0.093 | -1.136 | -1.288 |  |  |
|  | p |  | NS (0.926) | NS (0.256) | NS (0.198) |  |  |
| Or | T0 | 16 | 189 (138-412) | 144 (76-248) | 48 (0-211) | 7.84 | 0.020 |
|  | T1 | 11 | 240 (108-305) | 92 (35-201) | 0 (0-53) | 14.89 | <0.001 |
|  | Z |  | -0.052 | 1.507 | -1.183 |  |  |
|  | p |  | NS (0.959) | NS (0.132) | NS (0.237) |  |  |
| Presence or absence of the cortical and peripheral responses of the Cb-MEPs at T0 and T1 |  |  |  |  |  |  |  |
| Muscle | Timing | Categorical data | N of responses per category |  |  | χ<sup>2</sup> (4) | p |
|  |  |  | FCC5h/FCC6h-Mz | C5/C6-Cz | C3/C4-Cz |  |  |
| Oc | T0 | Present | 24 | 18 | 14 | 14.03 | 0.007 |
|  |  | Absent | 2 | 5 | 12 |  |  |
|  |  | Peripheral | 1 | 4 | 1 |  |  |
|  | T1 | Present | 17 | 9 | 12 | 7.38 | NS |
|  |  | Absent | 6 | 10 | 12 |  |  |
|  |  | Peripheral | 4 | 8 | 3 |  |  |
| Or | T0 | Present | 19 | 16 | 10 | 19.90 | <0.001 |
|  |  | Absent | 0 | 1 | 10 |  |  |
|  |  | Peripheral | 8 | 10 | 7 |  |  |
|  | T1 | Present | 15 | 11 | 6 | 11.82 | 0.019 |
|  |  | Absent | 2 | 3 | 10 |  |  |
|  |  | Peripheral | 10 | 13 | 11 |  |  |

Responses induced by FCC5h/FCC6h-Mz stimulation were significantly larger than those induced by C3/C4-Cz for both Oc (p<0.001 at T0; p=0.004 at T1, respectively) and Or (p=0.008 at T0; p=0.001 at T1, respectively); when compared with those induced by C5/C6-Cz stimulation, they were considerably larger for Oc, but the significance did not survive Bonferroni correction for multiple comparisons (p=0.033 at T0; p=0.034 at T1, respectively). Comparing responses induced by C3/C4-Cz and C5/C6-Cz stimulation, they were significantly larger for C5/C6-Cz stimulation only for Or at T1 (p=0.008) and markedly enlarged at T0 (p=0.020).

Responses ranked in categorical data (Cb-MEP PRESENT, ABSENT or PERIPHERAL, see Figure 2 for examples) were significantly differently distributed using the different montages (see the lower part of **Table 3**).

**Figure 2:**
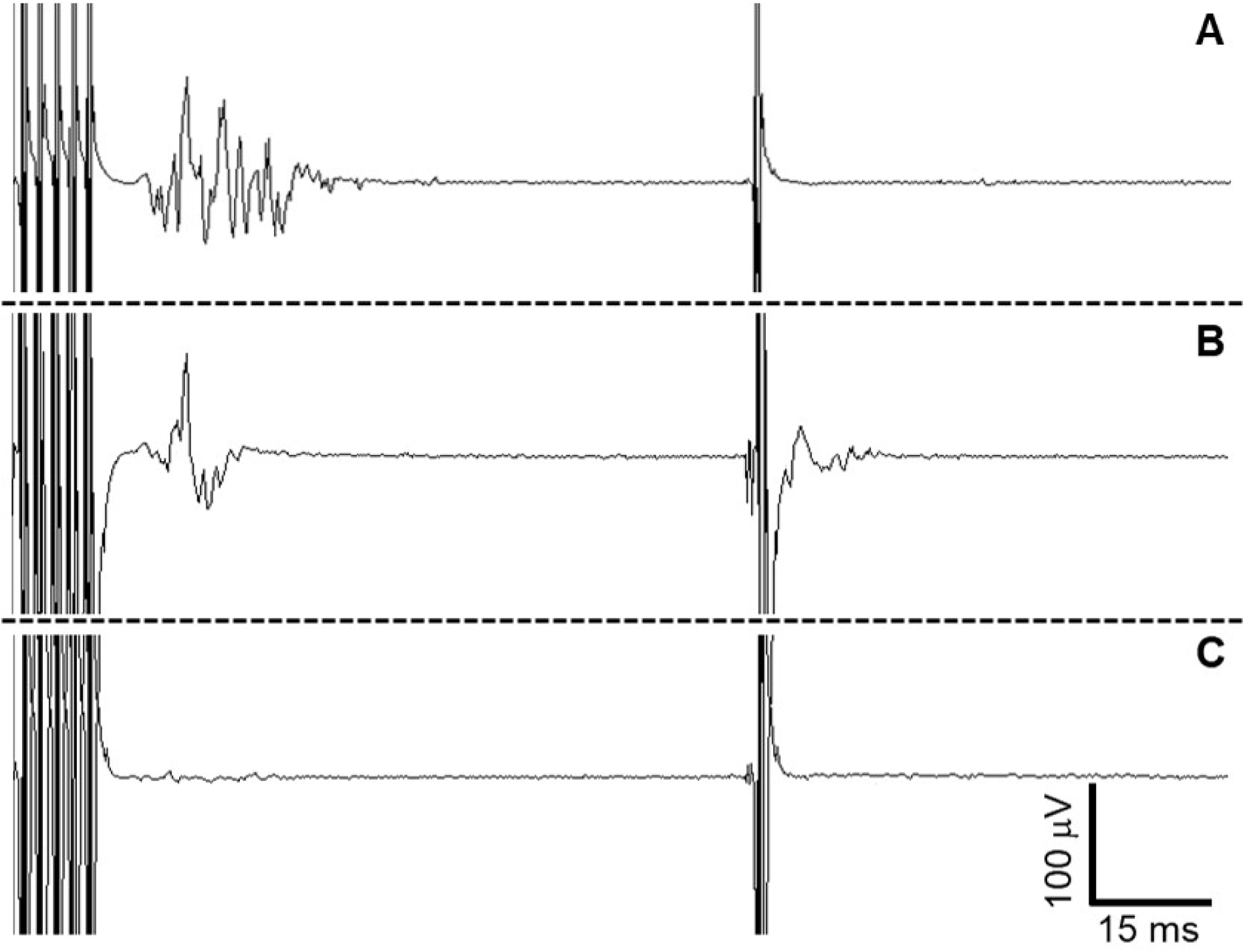
examples of Cb-MEP present, with peripheral responses and absent are shown in A (patient 1, T0), B (patient 6, T0) and C (patient 3, T1), respectively. Data shown refer to Cb-MEPs evoked with FCC5h/FCC6h-Mz stimulation and recorded from Orbicularis Oris.

### 3.4. Diagnostic measures

TES induced from FCC5h/FCC6h-Mz stimulation showed the higher accuracy (82.6%), specificity (88.9%), NPV and PPV (88.9% and 60.0%, respectively), while C5/C6-Cz stimulation had the higher sensitivity (66.7 %). Accuracy was 72.2% and 64.7% for TES from C5/C6-Cz and C3/C4-Cz, respectively.

Patients for whom a determination of true/false negative/positive was not applicable were 4 (13.3%), 9 (30.0%) and 10 (33.3%) for FCC5h/FCC6h-Mz, C5/C6-Cz and C3/C4-Cz stimulation, respectively.

The concordance of the three different TES-montages with postsurgical outcome in the single patients and the diagnostic measures of the three different TES-montages with respect to the postsurgical outcome are shown in **Table 4**.

**Table 4:** concordance of the three different TES-montages with postsurgical outcome in the single patients and diagnostic measures of the sample. FN=false negative; FP=false positive; TN=true negative; TP=true positive; CI=95% confidence interval; NPV=negative predictive value; PPV=positive predictive value; NA=not applicable.

| Patient | Postsurgical Outcome<br>(function of facial nerve) | Concordance |  |  |
| --- | --- | --- | --- | --- |
|  |  | FCC5h/FCC6h-Mz | C5/C6-Cz | C3/C4-Cz |
| #1 | WORSENING | NA | NA | NA |
| #2 | UNCHANGED | TN | NA | TN |
| #3 | WORSENING | TP | TP | TP |
| #4 | UNCHANGED | TN | FP | TN |
| #5 | WORSENING | TP | TP | NA |
| #6 | UNCHANGED | TN | TN | TN |
| #7 | UNCHANGED | FP | FP | FP |
| #8 | WORSENING | FN | FN | NA |
| #9 | UNCHANGED | TN | FP | TN |
| #10 | UNCHANGED | TN | TN | TN |
| #11 | UNCHANGED | FP | NA | NA |
| #12 | UNCHANGED | TN | NA | NA |
| #13 | UNCHANGED | TN | TN | TN |
| #14 | UNCHANGED | TN | TN | FP |
| #15 | UNCHANGED | TN | NA | TN |
| #16 | UNCHANGED | PATIENT EXCLUDED |  |  |
| #17 | UNCHANGED | TN | TN | FP |
| #18 | WORSENING | TP | FN | FN |
| #19 | UNCHANGED | TN | NA | TN |
| #20 | WORSENING | PATIENT EXCLUDED |  |  |
| #21 | WORSENING | NA | NA | NA |
| #22 | UNCHANGED | TN | TN | NA |
| #23 | WORSENING | NA | TP | NA |
| #24 | UNCHANGED | TN | NA | NA |
| #25 | WORSENING | FN | TP | FN |
| #26 | UNCHANGED | PATIENT EXCLUDED |  |  |
| #27 | UNCHANGED | NA | NA | NA |
| #28 | UNCHANGED | TN | TN | TN |
| #29 | UNCHANGED | TN | TN | TN |
| #30 | UNCHANGED | TN | TN | FP |
| Diagnostic measures |  |  |  |  |
| True Negative |  | 16 | 9 | 10 |
| True Positive |  | 3 | 4 | 1 |
| False Negative |  | 2 | 2 | 2 |
| False Positive |  | 2 | 3 | 4 |
| Not Applicable |  | 4 | 9 | 10 |
| Sensitivity, % (CI, %) |  | 60.0 (17.0-92.7) | 66.7 (24.1-94.0) | 33.3 (1.8-87.5) |
| Specificity, % (CI, %) |  | 88.9 (63.9-98.1) | 75.0 (42.8-93.3) | 71.4 (42.0-90.4) |
| NPV, % (CI, %) |  | 88.9 (63.9-98.1) | 81.8 (47.8-96.8) | 83.3 (50.9-97.1) |
| PPV, % (CI, %) |  | 60.0 (17.0-92.7) | 57.1 (20.2-88.2) | 20.0 (1.1-70.1) |
| Accuracy, % (CI, %) |  | 82.6 (67.1-98.1) | 72.2 (51.5-93.0) | 64.7 (42.0-89.8) |

## 4. Discussion

IONM helps the assessment of the integrity of the neural structures during surgical procedures. Since the first work of the group of Chang (Chang et al., 1999) to the study carried out on the huge sample of 100 patients made by Fernández-Conejero (Fernández-Conejero et al., 2022), the use of Cb-MEPs has become increasingly essential in monitoring the functioning of facial nerve during skull base surgery and/or in the brainstem surgery (Zhou and Kelly, 2001; Fukuda et al., 2008). To date, a general agreement of the usefulness of Cb-MEPs has been achieved, considering that it is able to determine the facial nerve outcome with a good accuracy (Akagami et al., 2005, Dong et al., 2005, Fukuda et al., 2008; Morota et al., 2010), despite some limitations due to the challenging interpretation if a response arises from peripheral or central stimulation (Tellez et al., 2016). C3/C4-Cz TES montage is the most diffuse to determine Cb-MEPs, although the São Paulo group (Verst et al., 2012; Verst et al., 2013) proposed the C5/C6-Cz stimulation, with similar effectiveness, and even in Dong et al., 2005, TES montages alternative to C3/C4-Cz were used. Hence, the debate moves progressively from demonstrating the usefulness of Cb-MEPs to finding the best technical specifications and procedures to limit pitfalls and increase the diagnostic yield of the technique.

### 4.1. Safety and feasibility of the FCC5h/FCC6h-Mz stimulation

First aim of this study was to prove safety and feasibility of FCC5h/FCC6h-Mz stimulation and to compare the Cb-MEPs data, especially the peak-to-peak amplitude, obtained from C3/C4-Cz, C5/C6-Cz and FCC5h/FCC6h-Mz montage during skull base surgery. We demonstrated the safety of FCC5h/FCC6h-Mz stimulation, given that no side effect was observed, such as seizures or after-discharges. Obviously, a larger number of cases are necessary to assess the actual risk of side effects, but our data are really promising.

With regard to the feasibility, we tried to obtain Cb-MEPs from FCC5h/FCC6h-Mz with the lowest voltage possible. In our sample, the mean voltage used was about 180 V, very similar to that used for C5/C6-Cz stimulation, but significantly lower than that used for C3/C4-Cz stimulation, in the work comparing different stimulation sites most similar to ours (Verst et al., 2012). Furthermore, the voltage used by Fukuda et al, in a study assessing facial nerve outcome with C3/C4-Cz stimulation performed in 2008, was higher than ours. The comparison between studies in which the intensity, not the voltage, of the current delivered was collected is more difficult (Fernández-Conejero et al., 2022), due to the unpredictable value of tissue resistance in the single cases.

Since the disappearance of the response, the threshold increase and the morphology simplification, beyond the amplitude reduction, have been considered criterion for MEP monitoring (see MacDonald DB, 2017, for an overview on this topic), the debate on which is the best among them still remains open; anyway for the surgery involving the brain, brainstem, and facial nerve monitoring, the reduction of the peak-to-peak amplitude greater than 50% is considered a major warning criterion based on several works (Dong et al., 2005; Neuloh et al., 2009; Verst et al., 2012; Verst et al., 2013; Fernández-Conejero et al., 2022). Hence, we considered the variations of the peak-to-peak amplitude to compare the Cb-MEPs obtained from the three montages studied: the peak-to-peak amplitude of Cb-MEPs obtained by FCC5h/FCC6h-Mz stimulation were significantly larger than those obtained by C3/C4-Cz and by C5/C6-Cz stimulation, for both Oc and Or and for Oc.

This result probably is related to the better position of the FCC5h/FCC6h-Cz dipole with respect to the cortical motor area of the face than the other two montages. The use of a low stimulus voltage is extremely important, in order to reduce the total amount of charge delivered, the risk of seizure and other complications (albeit overall low in skull base surgery) (MacDonald, 2002; Ulkatan et al., 2006). Again, with a low stimulus voltage the patient’s movements are smaller, thus reducing IONM interference with the surgical maneuvers and making the continuous monitoring easier.

Furthermore, the low stimulus voltage restrains the risk of leaking current (see also below, 4.2., analysis of the response category).

### 4.2. Analysis of the response category

In the second part of the work, we analyzed the response category (present, absent or peripheral) related to different stimulation montages. Remarkably, the FCC5h/FCC6h-Mz stimulation was more effective than others in inducing measurable Cb-MEPs, for both Oc and Or, while with C5/C6-Cz stimulation there was a higher risk of having a peripheral response and with C3/C4-Cz stimulation there was a higher probability of having no valuable Cb-MEPs (absent response), at the voltage used. If the advantage of FCC5h/FCC6h-Mz with respect to C3/C4-Cz stimulation is obvious, some considerations have to be accounted to underline why limiting the presence of a peripheral response is mandatory in monitoring the facial nerve functioning. It’s well-known that high or even moderate TES intensity can cause a leaking current activating the facial nerve itself, determining confounding peripheral responses (MacDonald et al., 2013). Hence, we considered a response as peripheral according to the suggestions of Deletis (Deletis et al., 2009b), i.e. when a single stimulus was sufficient to determine a measurable response. While keeping in mind the cautions of Tellez (Tellez et al., 2016), who demonstrated that in some cases a peripheral response emerges only when a train of stimuli was delivered (and not with a single stimulus), on the other hand the presence of a response following a single stimulus is almost invariably determined by a direct activation of facial nerve (Deletis et al., 2009b). In this line, the presence of a significant higher percentage of peripheral response can be considered representative of a higher risk of leaking currents activating facial nerve, when C5/C6-Cz stimulation is performed. As the whole, the data suggest that FCC5h/FCC6h-Mz montages allows to obtain Cb-MEPs useful for monitoring in a greater number of patients than other montages, at low voltage stimulation, even if the risk of some mistakes in the interpretation of the responses is not completely eliminated, and the most appropriate montage is not easily predictable in the single cases (see also below, 5, conclusions and practical considerations).

### 4.3. Analysis of the diagnostic measures

In the last part of our work, we assessed the diagnostic measures derived from the analysis of Cb-MEPs obtained with the three montages studied with respect to the clinical assessment of facial nerve after surgery.

Overall, considering the confidence intervals based on the size sample, our results cannot be conclusive, even if FCC5h/FCC6h-Mz stimulation showed the higher accuracy, specificity, NPV and PPV. Independently of montage used, our data confirm the reliability of the specificity and NPV of the Cb-MEPs (for all the TES used, higher than 70% and 80%, respectively), which means that the stability of the Cb-MEPs during and at the end of the intervention indicates that the facial nerve was spared (Dong et al., 2005; Fukuda et al., 2008; Fernández-Conejero et al., 2022). With respect to specificity, sensitivity achieved less important results, reaching 66% maximum with C5/C6-Cz stimulation. The high level of NPV, specificity and accuracy obtained with FCC5h/FCC6h-Mz stimulation was remarkable because obtained at low stimulus voltage.

The accuracies of the different stimulations included in our study are comparable with those of other studies (Akagami et al., 2005; Dong et al., 2005; Fukuda et al., 2008; Fernández-Conejero et al., 2022), even with some slight differences. Some of them can be explained by the difference in the etiologies of the lesions between studies, since in our sample acoustic neurinoma was present in about one third of the cases, while it was largely prevalent in the other studies. Again, we considered the early prognosis of facial nerve functioning, while the long-term prognosis was used in Dong et al., 2005: in some patients the complete recovery of functions can only occur after some time, for this reason the diagnostic measures post-operatively and once recovery is completed may be different. Moreover, also the threshold fixed at ≤ 50% of baseline to identify a significant decrease in the Cb-MEPs could have determined some little differences with other studies in which a Cb-MEP amplitude ≤ 35% of baseline was fixed as threshold (Dong et al., 2005; Fernández-Conejero et al., 2022). Probably these different options in the calculation of FN/FP/TN/TP are probably more theoretical than practical (the same Fernández-Conejero et al., 2022, alerted the surgeon when they detected a decrease of Cb-MEP amplitude of 50%), but they may have contributed in the slight variations between studies.

## 5. Conclusions and practical considerations

All our data converge in identifying FCC5h/FCC6h-Mz as the best TES montage for monitoring the facial nerve functioning: this montage allows maintaining excellent diagnostic accuracy with low stimulus voltages (intensities), while reducing the probability to obtain peripheral responses to transcranial stimulation.

However, it is not definite that in the single cases, such as for patients #13 and #18, where C3/C4-Cz and C5/C6-Cz showed better results, respectively. Hence, the best approach in the daily routine could be trying TES with different montages at baseline and choose “the best one”, based on the amplitude of the responses, their reproducibility and the absence of peripheral responses. This approach is not particularly time-consuming, lasting about 15-20 minutes, but it largely enhances the possibility of IONM, increasing its accuracy, especially when a low voltage TES is used.

## Author contributions

Mikael Gian Andrea Izzo: conceptualization (lead), project administration, writing-review & editing; Davide Rossi Sebastiano: conceptualization (supporting), formal analysis (supporting), visualization, writing-original draft (lead); Valentina Catanzaro: data-curation (lead-equal); Ylenia Melillo: data-curation (lead-equal); Ramona Togni: data-curation (lead-equal); Elisa Visani: formal analysis (lead); Jacopo Falco: writing-original draft (supporting); Cecilia Casali: writing-original draft (supporting); Marco Gemma: formal analysis (supporting); Paolo Ferroli: supervision (equal); Annamaria Gallone: data-curation (supporting); Daniele Cazzato: data-curation (supporting); Grazia Devigili: data-curation (supporting); Sara Alverà: data-curation (supporting); Paola Lanteri: supervision (equal).

## Data Availability

All data produced in the present study are available upon reasonable request to the authors

## List of abbreviations

BIS: Bispectral Index
CI: 95% Confidence Interval
Cb-MEPs: Corticobulbar-Motor Evoked Potentials
EEG: ElectroEncephalography
EMG: ElectroMyoGraphy
FN: False Negative
FP: False Positive
IONM: IntraOperative NeuroMonitoring
Oc: Orbicularis Oculi
Or: Orbicularis Oris
NA: Not Applicable
NPV: Negative Predictive Value
PPV: Positive Predictive Value
TES: Transcranial Electrical Stimulation
TN: True Negative
TP: True Positive

## Acknowledgments

We want to thank the Italian Ministry of Health which supported our study.

## References

Akagami R, Dong CC, Westerberg BD. Localized transcranial electrical motor evoked potentials for monitoring cranial nerves in cranial base surgery. Neurosurgery. 2005 Jul;57(1 Suppl):78–85; discussion 78-85. doi: 10.1227/01.neu.0000163486.93702.95. PMID: 15987572.

Avidan MS, Zhang L, Burnside BA, Finkel KJ, Searleman AC, Selvidge JA, Saager L, Turner MS, Rao S, Bottros M, Hantler C, Jacobsohn E, Evers AS. Anesthesia awareness and the bispectral index. N Engl J Med. 2008 Mar 13;358(11):1097–108. doi: 10.1056/NEJMoa0707361. PMID: 18337600.

Bossuyt PM, Reitsma JB, Bruns DE, Gatsonis CA, Glasziou PP, Irwig LM, Lijmer JG, Moher D, Rennie D, de Vet HC; STARD Group. Toward complete and accurate reporting of studies of diagnostic accuracy: the STARD initiative. Acad Radiol. 2003 Jun;10(6):664–9. doi: 10.1016/s1076-6332(03)80086-7. PMID: 12809421.

Chang SD, López JR, Steinberg GK. Intraoperative electrical stimulation for identification of cranial nerve nuclei. Muscle Nerve. 1999 Nov;22(11):1538–43. doi: 10.1002/(sici)1097-4598(199911)22:11<1538::aid-mus8>3.0.co;2-0. PMID: 10514231.

Deletis V, Urriza J, Ulkatan S, Fernandez-Conejero I, Lesser J, Misita D. The feasibility of recording blink reflexes under general anesthesia. Muscle Nerve. 2009 May;39(5):642–6. doi: 10.1002/mus.21257. PMID: 19347924. (a)

Deletis V, Fernandez-Conejero I, Ulkatan S, Costantino P. Methodology for intraoperatively eliciting motor evoked potentials in the vocal muscles by electrical stimulation of the corticobulbar tract. Clin Neurophysiol. 2009 Feb;120(2):336–41. doi: 10.1016/j.clinph.2008.11.013. Epub 2009 Jan 10. PMID: 19136297. (b)

Deletis V, Fernández-Conejero I, Ulkatan S, Rogić M, Carbó EL, Hiltzik D. Methodology for intra-operative recording of the corticobulbar motor evoked potentials from cricothyroid muscles. Clin Neurophysiol. 2011 Sep;122(9):1883–9. doi: 10.1016/j.clinph.2011.02.018. Epub 2011 Mar 25. PMID: 21440494.

Dickins JR, Graham SS. A comparison of facial nerve monitoring systems in cerebellopontine angle surgery. Am J Otol. 1991 Jan;12(1):1–6. PMID: 2012182.

Dong CC, Macdonald DB, Akagami R, Westerberg B, Alkhani A, Kanaan I, Hassounah M. Intraoperative facial motor evoked potential monitoring with transcranial electrical stimulation during skull base surgery. Clin Neurophysiol. 2005 Mar;116(3):588–96. doi: 10.1016/j.clinph.2004.09.013. PMID: 15721072.

Eichert N, Watkins KE, Mars RB, Petrides M. Morphological and functional variability in central and subcentral motor cortex of the human brain. Brain Struct Funct. 2021 Jan;226(1):263–279. doi: 10.1007/s00429-020-02180-w. Epub 2020 Dec 23. PMID: 33355695; PMCID: PMC7817568.

Fernández-Conejero I, Ulkatan S, Sen C, Miró Lladó J, Deletis V. Intraoperative monitoring of facial corticobulbar motor evoked potentials: Methodological improvement and analysis of 100 patients. Clin Neurophysiol. 2022 Oct;142:228–235. doi: 10.1016/j.clinph.2022.08.006. Epub 2022 Aug 24. PMID: 36081239.

Fukuda, M., Oishi, M., Takao, T., Saito, A., & Fujii, Y. (2008). Facial nerve motor-evoked potential monitoring during skull base surgery predicts facial nerve outcome. Journal of neurology, neurosurgery, and psychiatry, 79(9), 1066–1070.

Goto T, Muraoka H, Kodama K, Hara Y, Yako T, Hongo K. Intraoperative Monitoring of Motor Evoked Potential for the Facial Nerve Using a Cranial Peg-Screw Electrode and a “Threshold-level” Stimulation Method. Skull Base. 2010 Nov;20(6):429–34. doi: 10.1055/s-0030-1261270. PMID: 21772800; PMCID: PMC3134811.

Holmes NP, Tamè L, Beeching P, Medford M, Rakova M, Stuart A, Zeni S. Locating primary somatosensory cortex in human brain stimulation studies: experimental evidence. J Neurophysiol. 2019 Jan 1;121(1):336–344. doi: 10.1152/jn.00641.2018. Epub 2018 Dec 21. PMID: 30575432; PMCID: PMC6383658.

House JW, Brackmann DE. Facial nerve grading system. Otolaryngol Head Neck Surg. 1985 Apr;93(2):146–7. doi: 10.1177/019459988509300202. PMID: 3921901.

Isaacson B, Kileny PR, El-Kashlan HK. Prediction of long-term facial nerve outcomes with intraoperative nerve monitoring. Otol Neurotol. 2005 Mar;26(2):270–3. doi: 10.1097/00129492-200503000-00025. PMID: 15793418.

James ML, Husain AM. Brainstem auditory evoked potential monitoring: when is change in wave V significant? Neurology. 2005 Nov 22;65(10):1551–5. doi: 10.1212/01.wnl.0000184481.75412.2b. PMID: 16301480.

Kodama K, Kothbauer KF, Deletis V. Mapping and monitoring of brainstem surgery. Handb Clin Neurol. 2022;186:151–161. doi: 10.1016/B978-0-12-819826-1.00021-1. PMID: 35772884.

Lin VY, Houlden D, Bethune A, Nolan M, Pirouzmand F, Rowed D, Nedzelski JM, Chen JM. A novel method in predicting immediate postoperative facial nerve function post acoustic neuroma excision. Otol Neurotol. 2006 Oct;27(7):1017–22. doi: 10.1097/01.mao.0000235308.87689.35. PMID: 17006353.

MacDonald DB. Safety of intraoperative transcranial electrical stimulation motor evoked potential monitoring. J Clin Neurophysiol. 2002 Oct;19(5):416–29. doi: 10.1097/00004691-200210000-00005. PMID: 12477987.

Macdonald DB, Skinner S, Shils J, Yingling C; American Society of Neurophysiological Monitoring. Intraoperative motor evoked potential monitoring - a position statement by the American Society of Neurophysiological Monitoring. Clin Neurophysiol. 2013 Dec;124(12):2291–316. doi: 10.1016/j.clinph.2013.07.025. Epub 2013 Sep 18. PMID: 24055297.

MacDonald DB. Overview on Criteria for MEP Monitoring. J Clin Neurophysiol. 2017 Jan;34(1):4–11. doi: 10.1097/WNP.0000000000000302. PMID: 28045852.

Morota N, Ihara S, Deletis V. Intraoperative neurophysiology for surgery in and around the brainstem: role of brainstem mapping and corticobulbar tract motor-evoked potential monitoring. Childs Nerv Syst. 2010 Apr;26(4):513–21. doi: 10.1007/s00381-009-1080-7. Epub 2010 Feb 9. PMID: 20143075.

Neuloh G, Bogucki J, Schramm J. Intraoperative preservation of corticospinal function in the brainstem. J Neurol Neurosurg Psychiatry. 2009 Apr;80(4):417–22. doi: 10.1136/jnnp.2008.157792. Epub 2008 Dec 15. PMID: 19074927.

Oostenveld R, Praamstra P. The five percent electrode system for high-resolution EEG and ERP measurements. Clin Neurophysiol. 2001 Apr;112(4):713–9. doi: 10.1016/s1388-2457(00)00527-7. PMID: 11275545.

Rammeloo E, Schouten JW, Krikour K, Bos EM, Berger MS, Nahed BV, Vincent AJPE, Gerritsen JKW. Preoperative assessment of eloquence in neurosurgery: a systematic review. J Neurooncol. 2023 Dec;165(3):413–430. doi: 10.1007/s11060-023-04509-x. Epub 2023 Dec 14. PMID: 38095774.

Romstöck J, Strauss C, Fahlbusch R. Continuous electromyography monitoring of motor cranial nerves during cerebellopontine angle surgery. J Neurosurg. 2000 Oct;93(4):586–93. doi: 10.3171/jns.2000.93.4.0586. PMID: 11014536.

Shibasaki H. Human brain mapping: hemodynamic response and electrophysiology. Clin Neurophysiol. 2008 Apr;119(4):731–43. doi: 10.1016/j.clinph.2007.10.026. Epub 2008 Jan 9. PMID: 18187361.

Sollmann N, Krieg SM, Säisänen L, Julkunen P. Mapping of Motor Function with Neuronavigated Transcranial Magnetic Stimulation: A Review on Clinical Application in Brain Tumors and Methods for Ensuring Feasible Accuracy. Brain Sci. 2021 Jul 7;11(7):897. doi: 10.3390/brainsci11070897. PMID: 34356131; PMCID: PMC8305823.

Téllez MJ, Ulkatan S, Urriza J, Arranz-Arranz B, Deletis V. Neurophysiological mechanism of possibly confounding peripheral activation of the facial nerve during corticobulbar tract monitoring. Clin Neurophysiol. 2016 Feb;127(2):1710–1716. doi: 10.1016/j.clinph.2015.07.042. Epub 2015 Nov 10. PMID: 26564391.

Ulkatan S, Jaramillo AM, Téllez MJ, Kim J, Deletis V, Seidel K. Incidence of intraoperative seizures during motor evoked potential monitoring in a large cohort of patients undergoing different surgical procedures. J Neurosurg. 2017 Apr;126(4):1296–1302. doi: 10.3171/2016.4.JNS151264. Epub 2016 Jun 24. PMID: 27341047.

Verst SM, Chung TM, Sucena AC, Maldaun MV, Aguiar PH. Comparison between the C5 or C6-Cz electrode assembly and C3 or C4-Cz assembly for transcranial electric motor activation of muscular response of the contralateral facial nerve. Acta Neurochir (Wien). 2012 Dec;154(12):2229–35. doi: 10.1007/s00701-012-1505-z. Epub 2012 Oct 4. PMID: 23053280.

Verst SM, Sucena AC, Maldaun MV, Aguiar PH. Effectiveness of C5 or C6-Cz assembly in predicting immediate post operative facial nerve deficit. Acta Neurochir (Wien). 2013 Oct;155(10):1863–9. doi: 10.1007/s00701-013-1806-x. Epub 2013 Jul 18. PMID: 23864399.

Voets NL, Bartsch AJ, Plaha P. Functional MRI applications for intra-axial brain tumours: uses and nuances in surgical practise. Br J Neurosurg. 2022 Sep 23:1–16. doi: 10.1080/02688697.2022.2123893. Epub ahead of print. PMID: 36148501.

Youssef AS, Downes AE. Intraoperative neurophysiological monitoring in vestibular schwannoma surgery: advances and clinical implications. Neurosurg Focus. 2009 Oct;27(4):E9. doi: 10.3171/2009.8.FOCUS09144. PMID: 19795957.

Zhou HH, Kelly PJ. Transcranial electrical motor evoked potential monitoring for brain tumor resection. Neurosurgery. 2001 May;48(5):1075–80; discussion 1080-1. doi: 10.1097/00006123-200105000-00021. PMID: 11334274.

